# Surrogate markers of metabolic syndrome (MetS) and insulin resistance (IR) in children and young adults with type 1 diabetes: A systematic review & meta-analysis

**DOI:** 10.1101/2023.05.23.23290372

**Authors:** Sukeshini Khandagale, Vinesh Kamble, Chirantap Oza, Shital Bhor, Anuradha Khadilkar, Satyajeet Khare

**Affiliations:** Symbiosis School of Biological Sciences, Symbiosis International University, Pune 412115; Hirabai Cowasji Jehangir Medical Research Institute, Jehangir Hospital, Pune 411001

**Keywords:** Biomarkers, insulin resistance, metabolic syndrome, type 1 diabetes, children

## Abstract

**Introduction:** Metabolic syndrome (MetS), a collection of risk factors for cardiovascular disease, and Insulin resistance (IR) are associated with diabetes. The diagnosis of both these conditions are based on specific clinical parameters. However, the efficacy of these parameters has not been systematically studied in paediatric population with T1DM.

**Methodology:** We performed a systematic review and meta-analysis in paediatric populations of type 1 diabetes. We assessed the strength of association of the parameters of MetS and IR. The meta-analysis was performed using ‘metaphor’ package in R. A random effect model was used to study the strength of association by estimating Hedge’s g.

**Results:** The systematic review resulted in identification of 30 studies on MetS and IR in paediatric patients with T1DM. Insulin dosage and HbA1C, markers for glycaemic condition showed no association with MetS in patients with T1DM. In the lipid profile, increased triglyceride (TG) and low-density lipoprotein (LDL) showed better effect size than reduced high-density lipoprotein (HDL). In case of insulin resistance, heterogeneous nature of studies made it difficult to carry out a meta-analysis. A descriptive review of existing and novel markers is thus provided.

**Conclusion:** Lack of association between markers of glycaemic condition suggested that MetS may develop independently of glycaemic control in children with T1DM. Other than TG and HDL, LDL may be used in the diagnosis of MetS. From the descriptive analysis it could be observed that a standard protocol for the diagnosis of IR is needed.

**Research highlight:**

1. The systematic review identified 30 research articles on the markers of MetS and IR in children with T1DM
2. Markers of glycaemic control did not associate with MetS in children with T1DM
3. Low Density Lipoprotein (LDL) levels showed a strong effect on MetS in children with T1DM suggesting it’s application as a parameter for the diagnosis of MetS.

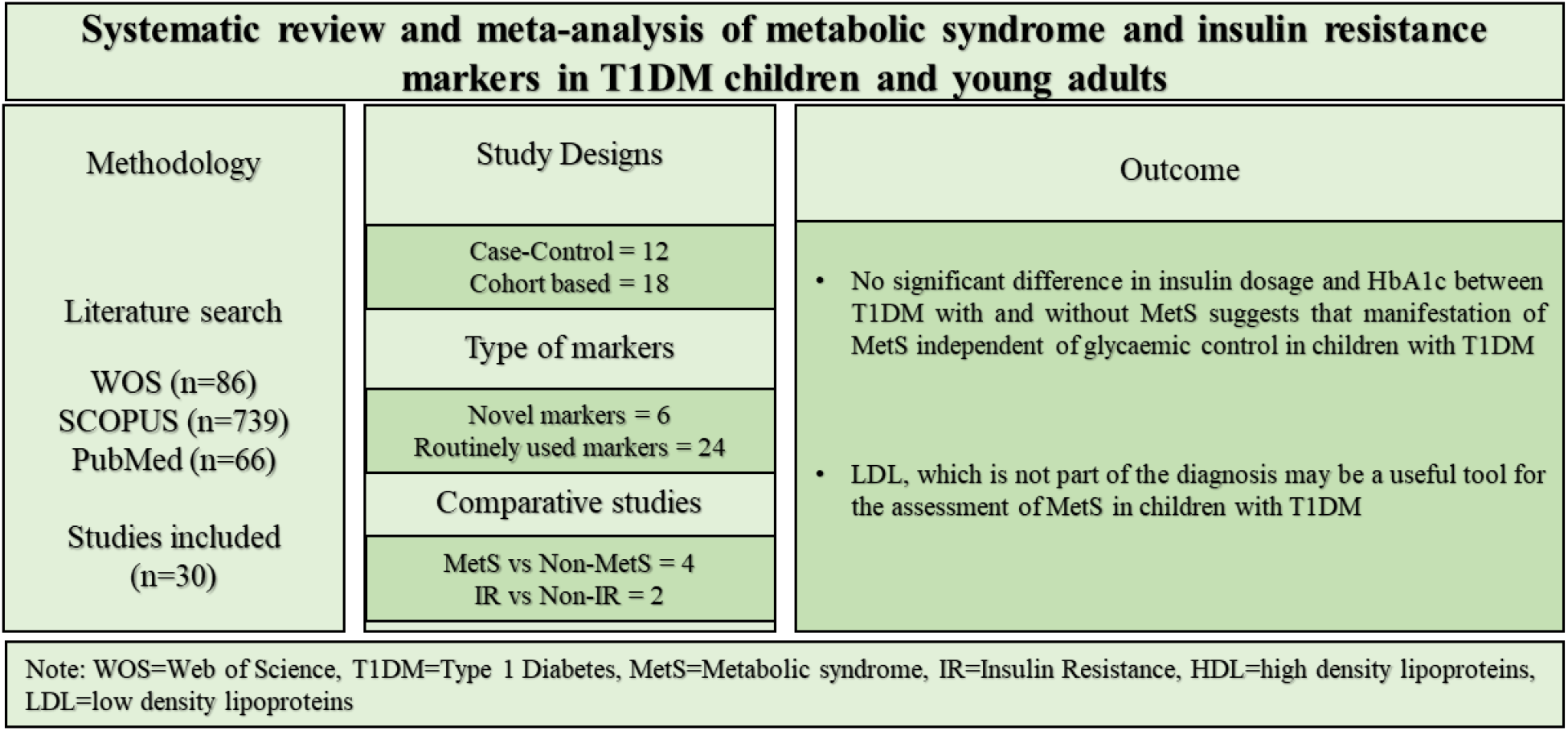

## Introduction

Type 1 diabetes mellitus (T1DM) is an autoimmune disorder in which the pancreatic beta cells are destroyed by the host immune system. The patient, as a result, is treated with exogenous insulin. The peak age for T1DM diagnosis is 5-9yrs and 10-14yrs^1,2^. The prevalence of T1DM is rising among the young population^3–6^. Patient with recently diagnosed T1DM generally have a lower body mass index; however, prevalence of obesity has increased among these patients during the recent decades^7^. Obesity is associated with insulin resistance and metabolic syndrome which are the risk factors for cardiovascular diseases. Therefore, diagnosis and management of metabolic syndrome (MetS) and insulin resistance (IR) are crucial for the prevention of cardiometabolic risks.

The prevalence of MetS in T1DM is suggested to be 23.7% and is increasing^8,9^. The diagnosis of MetS is based on three different criteria that are laid down by the World health Organization (WHO), the National Cholesterol Education Programme Adult Treatment Panel III (NCEP ATP III), and the International Diabetes Federation (IDF)^10^. These criteria are based on anthropometric measurements such as waist circumference (WC), hypertension (HTN) and biochemical parameters such as the lipid profile (Table 1).

**Table 1:** Diagnostic criteria for metabolic syndrome according to different organization.

| Criteria laid by | Components of MetS | Cut-offs for adults |
| --- | --- | --- |
| WHO (1998)<sup>11</sup> | Insulin resistance (by impaired fasting glucose (FG) or impaired glucose tolerance (IGT) or Hyperinsulinemic Euglycaemic Clamp (HEC)) with (any 2 of the following: obesity, dyslipidemia, high systolic, high diastolic blood pressure, increased urine microalbuminuria) | FG>100mg/dl, IGT>140 mg/dl 120 minutes after ingestion of 75 grams of glucose, WHR: $\geq 0.90$(M), $0.85$(F) or BMI$\geq 30$kg/m<sup>2</sup>, TG$\geq 150$mg/dl or HDL-C$\leq 35$mg/dl(M), $39$mg/dl(F), BP$\geq 160/90$mmHg, Urinary albumin excretion of 20 μg/min or albumin-to-creatinine ratio of 30 mg/g |
| NCEP ATP III (2005 revised)<sup>10</sup> | Any 3 of five: Obesity, Hyperglycemia, dyslipidemia, high systolic or high diastolic BP. | Waist circumference: $\geq 40$ inches (M), $\geq 35$ inches (F), Fasting glucose $\geq 100$ mg/dl or Rx, TG$\geq 150$ mg/dl or Rx, HDL cholesterol$\leq 40$ mg/dl (M), $\leq 50$ mg/dl (F); or Rx, HTN$\geq 130$ mmHg systolic or $\geq 85$ diastolic or Rx. |
| IDF(2007)<sup>12</sup> | Central obesity (by waist circumference) with (2 of the four criteria: FG, TG, HDL, BP)<br>*If BMI$>30$kg/m<sup>2</sup> central obesity can be assumed | Waist circumference: $\geq 94$cm(M), $\geq 80$cm(F), FG$\geq 100$mg/dl, TG$\geq 150$mg/dl, HDL$\leq 40$mg/dl(M), $\leq 50$mg/dl(F), BP$\geq 130$mmHg or $\geq 85$mmHg diastolic or Rx |

**Note:** WHR (waist to hip ratio), HDL (High Density Lipoprotein), BP (Blood Pressure), BMI (Body Mass Index), Rx (on drugs of treatment for the condition)

Most of the cut-offs for the diagnosis of MetS are developed for the adult population^13–15^. These parameters are modified only by changing the threshold for use in paediatric population. In case of type 1 diabetes, these MetS parameters may not be stable due to exogenous insulin therapy and pubertal age of the paediatric patients.

Along with MetS, an increased prevalence of insulin resistance (IR) is also observed in patients with T1DM. Insulin resistance is generally a characteristic of T2DM and therefore, the development of insulin resistance in T1DM is termed as double diabetes^16–20^. Various factors such as food habits, reduced physical activity, gender, age, and genetic predisposition may have a role to play in the development of IR in T1DM ^21,22^. IR has been associated with various other disorders such as polycystic ovary syndrome (PCOS)^23,24^, non-alcoholic fatty liver disease (NAFLD)^25–27^ etc. The increased frequency of IR in T1DM makes children with T1DM more prone to the development of PCOS and NAFLD. Presence of IR in children with T1DM also increases the risk of development of various microvascular complications^28^. Hence, the diagnosis of IR may help clinicians to implement preventive measures or add an adjuvant treatment.

The diagnosis of IR in T2DM depends on measurement of fasting insulin levels which are negligible in T1DM. Therefore, the indices used for the diagnosis of IR in Type 2 diabetes have little use in T1DM. The gold standard method for the diagnosis of IR in children with T1DM is the Hyperinsulinemia Euglycemic Clamp (HEC) in which the glucose concentration is maintained by variable infusion of exogenous glucose and insulin^29^. However, the HEC technique is expensive, and space and time consuming. Therefore, various alternate methods have been developed for the diagnosis of IR that rely on indirect markers such as estimated glucose disposal rate (eGDR)^30–33^, Insulin Sensitivity Score (ISS)^34^ and, insulin sensitivity equation (eIS)^35^. The indices for the diagnosis of IR in

T1DM are provided by Epidemiology of Diabetes Complications (EDC), Search for diabetes in youth (SEARCH), and Coronary Artery Calcification in T1DM (CACTI) (Table 2).

**Table 2:** Indices provided for calculation of insulin resistance in Type 1 diabetes.

| Groups | Equations | Target population | Threshold for IR |
| --- | --- | --- | --- |
| IDF (2007)<sup>36</sup> | $eGDR = 24.31 - 12.22 \times (WHR) - 3.29 \times (Hypertention) - 0.57 \times (A1C[\%])$ | Adult T1DM (compared with HEC) | Does not provide cut-offs, usually studied by dividing groups in tertiles or quartiles<sup>37</sup>. |
| SEARCH (2011)<sup>34,38</sup> | $IS \text{ scores} = \text{Exp}(4.64725 - 0.02032(\text{waist[cm]}) - 0.09779(\text{HbA1c[\%]}) - 0.00235(\text{TG[mg/dL]}))$ | Adolescence with T1DM, T2DM and non-diabetic (compared with HEC) | Does not have provide cut-offs |
| CACTI (2011)<sup>39</sup> | $eIS = \text{Exp}(4.1075 - 0.01299 \times (\text{waist[cm]}) - 1.05819 \times (\text{insulin dose}) - 0.00354 \times (\text{TG[mg/dL]}) - 0.00802 \times (\text{DBP[mmHg]})$ | Adult T1DM (compared with HEC) | Does not provide cut-offs, parameters are tested as models with and without adiponectin and fasting/non-fasting state |

**Note:** EDC (Epidemiology of Diabetes Complications), SEARCH (SEARCH for Diabetes in Youth), CACTI (Coronary Artery Calcification in T1DM), eGDR (estimated glucose disposal rate), IS (insulin sensitivity score), eIS (estimated insulin sensitivity, WHR (waist to hip ratio), TG(Triglycerides), DBP (Diastolic Blood Pressure).

The indices for IR in T1DM have been validated by direct comparison with HEC^34,38,40,39^. There are no threshold or cut-offs provided for these indices. However, many authors have provided cohort based thresholds. Most of these studies include adults with T1DM. The study by Oza et al has provided cut-offs for all three indices in children with T1DM (Supplementary Table 1). The exogenous insulin administration and pubertal age may interfere with the existing parameters of MetS and IR. Therefore, a systematic review and meta-analysis is needed for both these conditions.

In this systematic review we have attempted to summarize and compare various individual indirect markers for the diagnosis of MetS and IR in T1DM for their efficacy. Our analysis suggests that metabolic syndrome may develop independently irrespective of glycaemic control in children with T1DM. In lipid profile, LDL and Triglycerides were found to exert a strong effect on metabolic syndrome whereas HDL exerted a moderate effect. Lack of consistency in study designs for the detection of insulin resistance in T1DM made it difficult to compare the effect size of markers of IR. Therefore, we provide a descriptive review of the existing and novel markers for IR in T1DM.

### Methodology

This is an exploratory meta-analysis and follows the PRISMA (Preferred Reporting of Systematic Review and Meta-analysis) guidelines.

### Search strategy, and Inclusion and exclusion criteria

Two authors (SK and VK) independently searched for the relevant keywords in three databases (PubMed, SCOPUS, Web of Science) for identification of research articles related to metabolic syndrome and insulin resistance in children, adolescents, and young adults with T1DM. The search was performed till May 5, 2023.

The articles were from 1982 to 2023. The search for the relevant keywords was as follows (((“Type 1 Diabetes” OR “IDDM” OR “insulin dependent diabetes” OR “T1DM”) AND (“insulin resistance” OR “IR” OR “Metabolic syndrome” OR “MetS” OR “insulin sensitivity” OR “IS”)) AND (“Molecular markers” OR “markers” OR “Biological markers” OR “Clinical markers” OR “gene expression markers”)) AND (“Paediatric” OR “child” OR “children” OR “adolescent” OR “adolescence” OR “young adult”).

The search was limited to peer reviewed English articles. Only original research articles were included for this review. Studies that had a type 1 diabetes population with the age group <25yrs were retained. The studies were then imported to a Rayyan software for screening and removal of duplicates^41^. Studies using animal models, cell lines, and organ tissue samples were excluded. Studies including patients with complications of diabetes and on treatment other than insulin therapy were excluded.

### Selection of studies and data extraction

We segregated the studies based on presence or absence of metabolic syndrome and insulin resistance in the T1DM population. The studies that provide markers for such conditions, either standard (insulin dose, eGDR for IR, IDF criteria for MetS) or surrogate (BMI, WC etc.), were included in this review. Meta-analysis was performed only if multiple studies with similar parameters were available. Other studies were utilized for descriptive review. Parameters such as duration of diabetes, insulin dosage, HbA1c, and lipid profile were assessed in each study. The sample size, mean, and standard deviation (sd) for each parameter were recorded accordingly. If median and interquartile range were provided they were converted to estimated mean and variance depending on sample size^42^. Author names, publication year, ethnicity, and gender details of the population were also recorded for the studies that were part of the systematic review (Table 3).

**Table 3:**
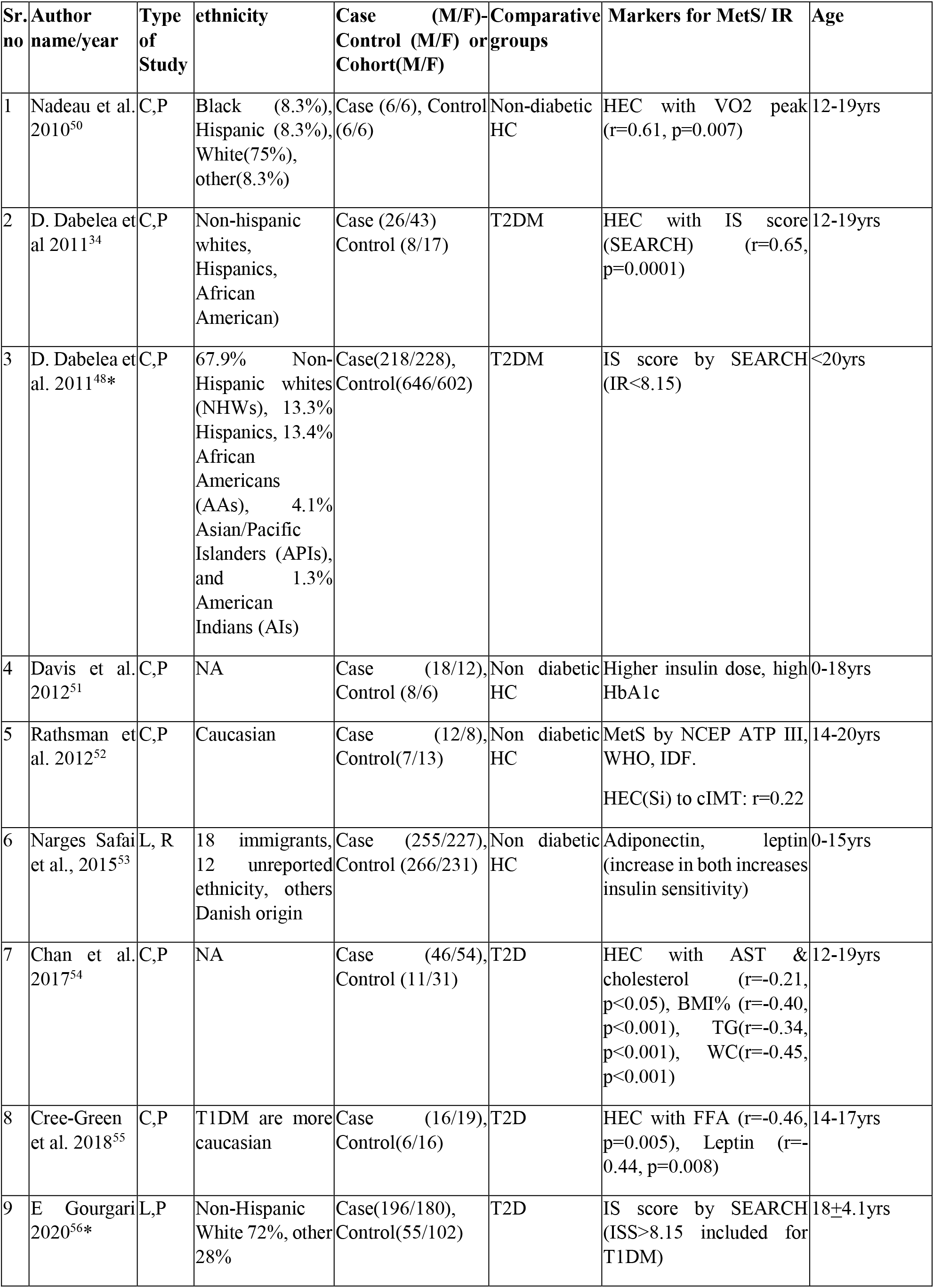
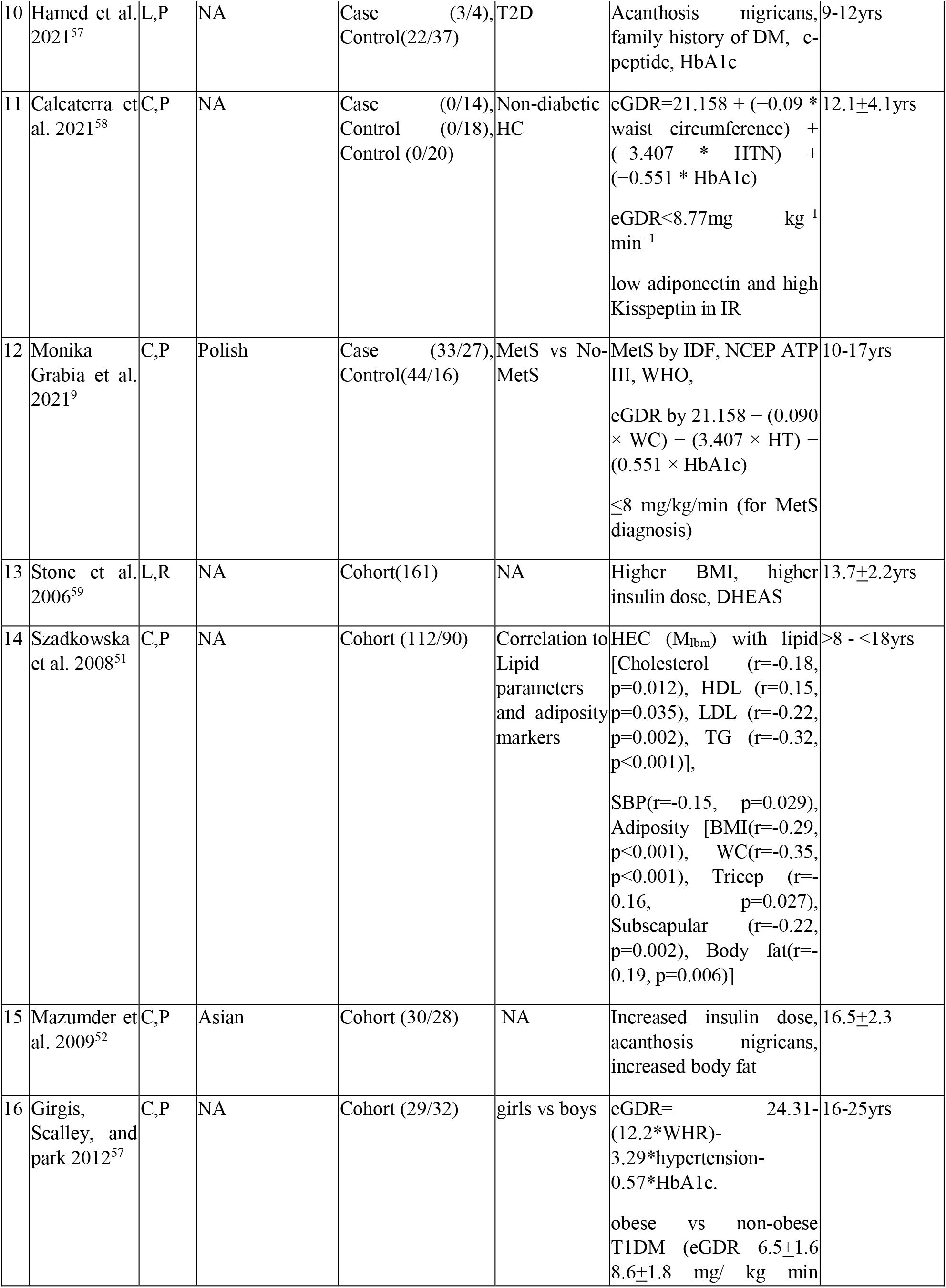
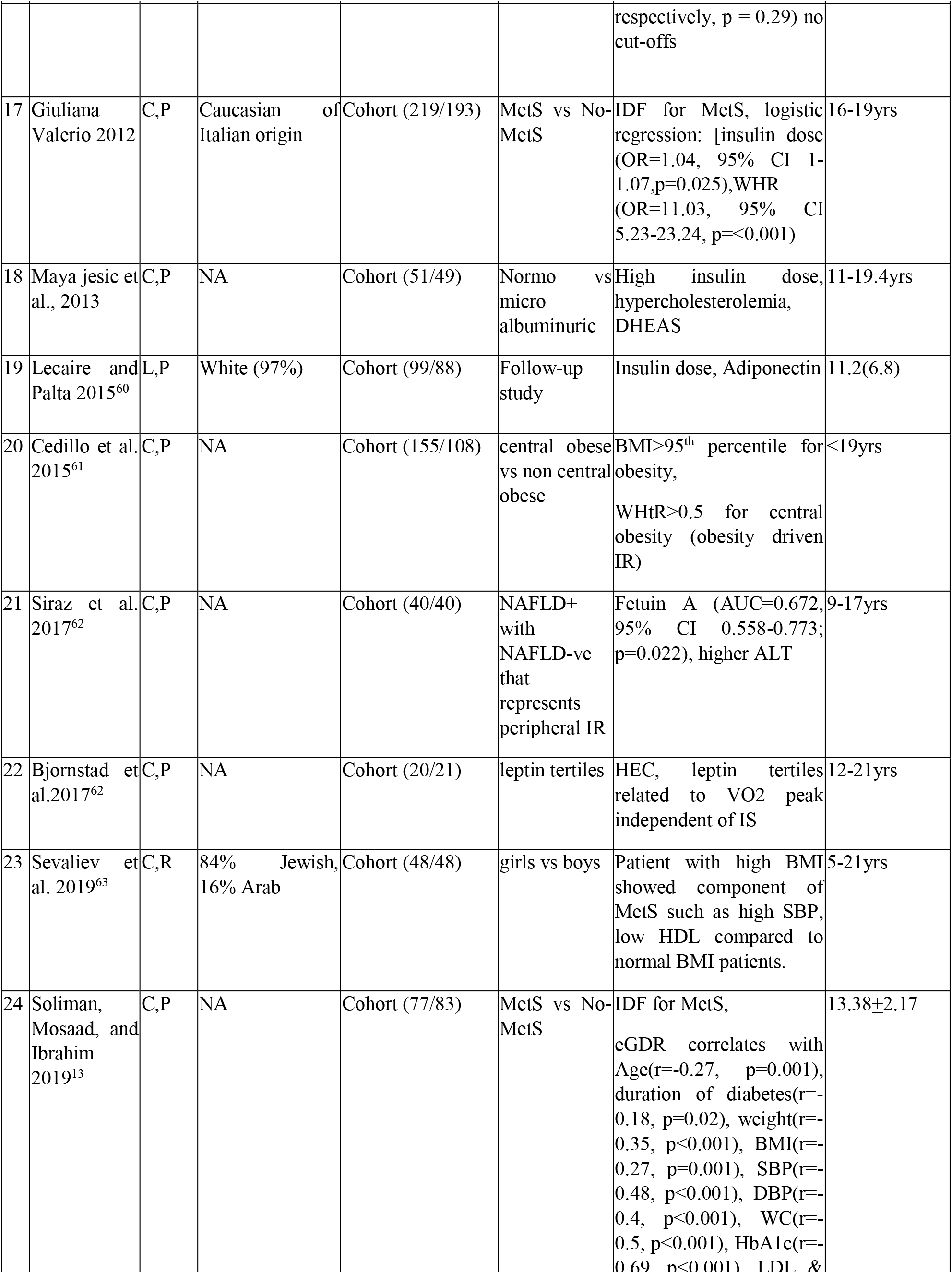
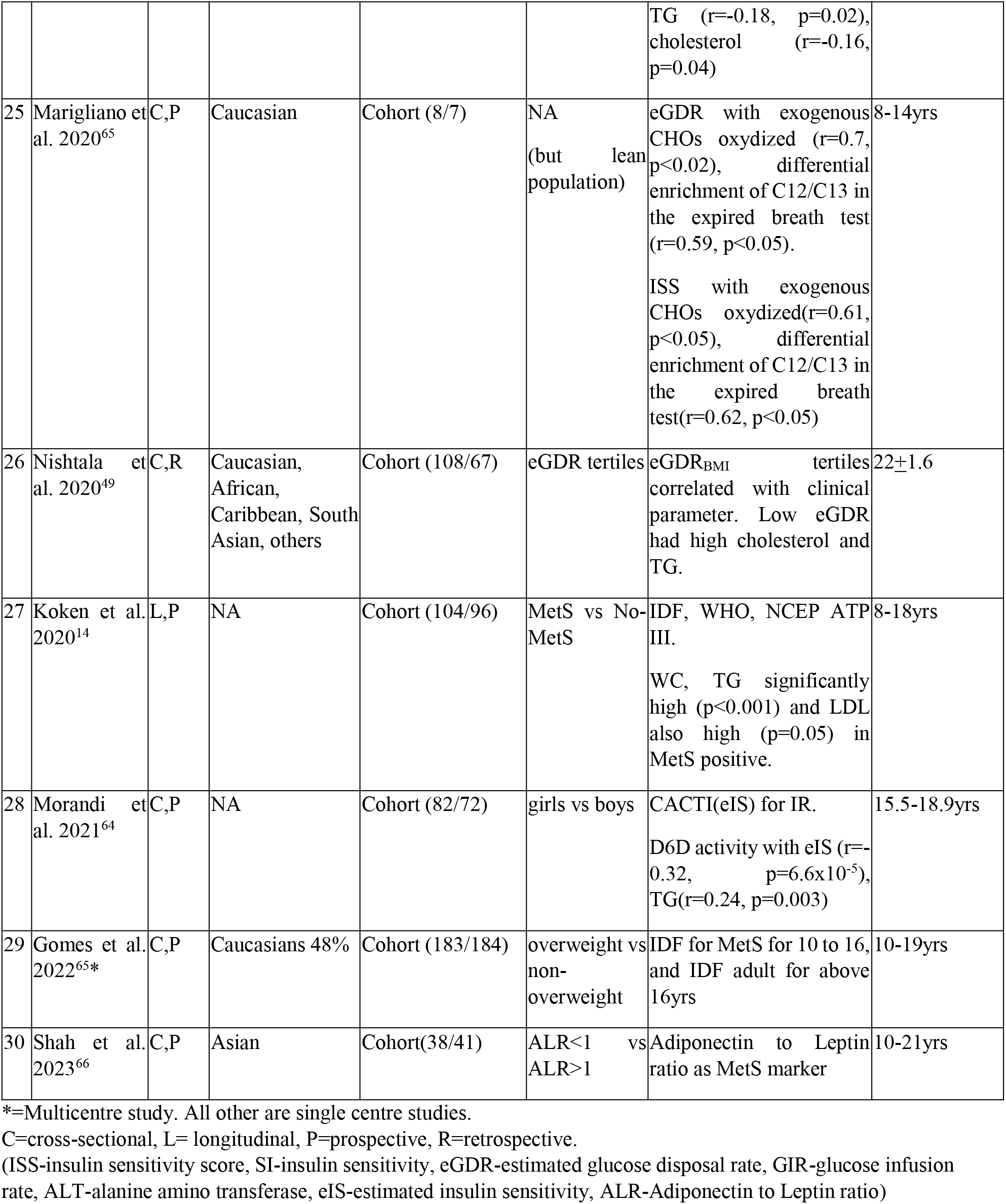
Characteristic of studies included in the systematic review

| Sr. no | Author name/year | Type of Study | ethnicity | Case (M/F)-Control (M/F) or Cohort(M/F) | Comparative groups | Markers for MetS/ IR | Age |
| --- | --- | --- | --- | --- | --- | --- | --- |
| 1 | Nadeau et al. 2010<sup>50</sup> | C,P | Black (8.3%), Hispanic (8.3%), White(75%), other(8.3%) | Case (6/6), Control (6/6) | Non-diabetic HC | HEC with VO2 peak (r=0.61, p=0.007) | 12-19yrs |
| 2 | D. Dabelea et al 2011<sup>34</sup> | C,P | Non-hispanic whites, Hispanics, African American) | Case (26/43) Control (8/17) | T2DM | HEC with IS score (SEARCH) (r=0.65, p=0.0001) | 12-19yrs |
| 3 | D. Dabelea et al. 2011<sup>48*</sup> | C,P | 67.9% Non-Hispanic whites (NHWs), 13.3% Hispanics, 13.4% African Americans (AAs), 4.1% Asian/Pacific Islanders (APIs), and 1.3% American Indians (AIs) | Case(218/228), Control(646/602) | T2DM | IS score by SEARCH (IR<8.15) | <20yrs |
| 4 | Davis et al. 2012<sup>51</sup> | C,P | NA | Case (18/12), Control (8/6) | Non diabetic HC | Higher insulin dose, high HbA1c | 0-18yrs |
| 5 | Rathsman et al. 2012<sup>52</sup> | C,P | Caucasian | Case (12/8), Control(7/13) | Non diabetic HC | MetS by NCEP ATP III, WHO, IDF.<br>HEC(Si) to cIMT: r=0.22 | 14-20yrs |
| 6 | Narges Safai et al., 2015<sup>53</sup> | L, R | 18 immigrants, 12 unreported ethnicity, others Danish origin | Case (255/227), Control (266/231) | Non diabetic HC | Adiponectin, leptin (increase in both increases insulin sensitivity) | 0-15yrs |
| 7 | Chan et al. 2017<sup>54</sup> | C,P | NA | Case (46/54), Control (11/31) | T2D | HEC with AST & cholesterol (r=-0.21, p<0.05), BMI% (r=-0.40, p<0.001), TG(r=-0.34, p<0.001), WC(r=-0.45, p<0.001) | 12-19yrs |
| 8 | Cree-Green et al. 2018<sup>55</sup> | C,P | T1DM are more caucasian | Case (16/19), Control(6/16) | T2D | HEC with FFA (r=-0.46, p=0.005), Leptin (r=-0.44, p=0.008) | 14-17yrs |
| 9 | E Gourgari 2020<sup>56*</sup> | L,P | Non-Hispanic White 72%, other 28% | Case(196/180), Control(55/102) | T2D | IS score by SEARCH (ISS>8.15 included for T1DM) | 18±4.1yrs |
| 10 | Hamed et al. 2021<sup>57</sup> | L,P | NA | Case (3/4), Control(22/37) | T2D | Acanthosis nigricans, family history of DM, c-peptide, HbA1c | 9-12yrs |
| 11 | Calcaterra et al. 2021<sup>58</sup> | C,P | NA | Case (0/14), Control (0/18), Control (0/20) | Non-diabetic HC | eGDR=21.158 + (-0.09 * waist circumference) + (-3.407 * HTN) + (-0.551 * HbA1c)<br>eGDR<8.77mg kg<sup>-1</sup> min<sup>-1</sup><br>low adiponectin and high Kisspeptin in IR | 12.1±4.1yrs |
| 12 | Monika Grabia et al. 2021<sup>9</sup> | C,P | Polish | Case (33/27), Control(44/16) | MetS vs No-MetS | MetS by IDF, NCEP ATP III, WHO,<br>eGDR by 21.158 - (0.090 × WC) - (3.407 × HT) - (0.551 × HbA1c)<br>≤8 mg/kg/min (for MetS diagnosis) | 10-17yrs |
| 13 | Stone et al. 2006<sup>59</sup> | L,R | NA | Cohort(161) | NA | Higher BMI, higher insulin dose, DHEAS | 13.7±2.2yrs |
| 14 | Szadkowska et al. 2008<sup>51</sup> | C,P | NA | Cohort (112/90) | Correlation to Lipid parameters and adiposity markers | HEC (M<sub>lbm</sub>) with lipid [Cholesterol (r=-0.18, p=0.012), HDL (r=0.15, p=0.035), LDL (r=-0.22, p=0.002), TG (r=-0.32, p<0.001)],<br>SBP(r=-0.15, p=0.029), Adiposity [BMI(r=-0.29, p<0.001), WC(r=-0.35, p<0.001), Tricep (r=-0.16, p=0.027), Subscapular (r=-0.22, p=0.002), Body fat(r=-0.19, p=0.006)] | >8 - <18yrs |
| 15 | Mazumder et al. 2009<sup>52</sup> | C,P | Asian | Cohort (30/28) | NA | Increased insulin dose, acanthosis nigricans, increased body fat | 16.5±2.3 |
| 16 | Girgis, Scalley, and park 2012<sup>57</sup> | C,P | NA | Cohort (29/32) | girls vs boys | eGDR= 24.31-(12.2*WHR)-3.29*hypertension-0.57*HbA1c.<br>obese vs non-obese T1DM (eGDR 6.5±1.6 8.6±1.8 mg/ kg min respectively, p = 0.29) no cut-offs | 16-25yrs |
| 17 | Giuliana Valerio 2012 | C,P | Caucasian of Italian origin | Cohort (219/193) | MetS vs No-MetS | IDF for MetS, logistic regression: [insulin dose (OR=1.04, 95% CI 1-1.07,p=0.025), WHR (OR=11.03, 95% CI 5.23-23.24, p=<0.001) | 16-19yrs |
| 18 | Maya jesic et al., 2013 | C,P | NA | Cohort (51/49) | Normo vs micro albuminuric | High insulin dose, hypercholesterolemia, DHEAS | 11-19.4yrs |
| 19 | Lecaire and Palta 2015<sup>60</sup> | L,P | White (97%) | Cohort (99/88) | Follow-up study | Insulin dose, Adiponectin | 11.2(6.8) |
| 20 | Cedillo et al. 2015<sup>61</sup> | C,P | NA | Cohort (155/108) | central obese vs non central obese | BMI>95<sup>th</sup> percentile for obesity, WHtR>0.5 for central obesity (obesity driven IR) | <19yrs |
| 21 | Siraz et al. 2017<sup>62</sup> | C,P | NA | Cohort (40/40) | NAFLD+ with NAFLD-ve that represents peripheral IR | Fetuin A (AUC=0.672, 95% CI 0.558-0.773; p=0.022), higher ALT | 9-17yrs |
| 22 | Bjornstad et al. 2017<sup>62</sup> | C,P | NA | Cohort (20/21) | leptin tertiles | HEC, leptin tertiles related to VO2 peak independent of IS | 12-21yrs |
| 23 | Sevaliev et al. 2019<sup>63</sup> | C,R | 84% Jewish, 16% Arab | Cohort (48/48) | girls vs boys | Patient with high BMI showed component of MetS such as high SBP, low HDL compared to normal BMI patients. | 5-21yrs |
| 24 | Soliman, Mosaad, and Ibrahim 2019<sup>13</sup> | C,P | NA | Cohort (77/83) | MetS vs No-MetS | IDF for MetS, eGDR correlates with Age(r=-0.27, p=0.001), duration of diabetes(r=-0.18, p=0.02), weight(r=-0.35, p<0.001), BMI(r=-0.27, p=0.001), SBP(r=-0.48, p<0.001), DBP(r=-0.4, p<0.001), WC(r=-0.5, p<0.001), HbA1c(r=-0.69, p<0.001), LDL & | 13.38±2.17 |
|  |  |  |  |  |  | TG (r=-0.18, p=0.02), cholesterol (r=-0.16, p=0.04) |  |
| 25 | Marigliano et al. 2020<sup>65</sup> | C,P | Caucasian | Cohort (8/7) | NA (but lean population) | eGDR with exogenous CHOs oxydized (r=0.7, p<0.02), differential enrichment of C12/C13 in the expired breath test (r=0.59, p<0.05).<br>ISS with exogenous CHOs oxydized(r=0.61, p<0.05), differential enrichment of C12/C13 in the expired breath test(r=0.62, p<0.05) | 8-14yrs |
| 26 | Nishtala et al. 2020<sup>49</sup> | C,R | Caucasian, African, Caribbean, South Asian, others | Cohort (108/67) | eGDR tertiles | eGDR<sub>BMI</sub> tertiles correlated with clinical parameter. Low eGDR had high cholesterol and TG. | 22±1.6 |
| 27 | Koken et al. 2020<sup>14</sup> | L,P | NA | Cohort (104/96) | MetS vs No-MetS | IDF, WHO, NCEP ATP III.<br>WC, TG significantly high (p<0.001) and LDL also high (p=0.05) in MetS positive. | 8-18yrs |
| 28 | Morandi et al. 2021<sup>64</sup> | C,P | NA | Cohort (82/72) | girls vs boys | CACTI(eIS) for IR.<br>D6D activity with eIS (r=-0.32, p=6.6x10<sup>-5</sup>), TG(r=0.24, p=0.003) | 15.5-18.9yrs |
| 29 | Gomes et al. 2022<sup>65*</sup> | C,P | Caucasians 48% | Cohort (183/184) | overweight vs non-overweight | IDF for MetS for 10 to 16, and IDF adult for above 16yrs | 10-19yrs |
| 30 | Shah et al. 2023<sup>66</sup> | C,P | Asian | Cohort(38/41) | ALR<1 vs ALR>1 | Adiponectin to Leptin ratio as MetS marker | 10-21yrs |

\*=Multicentre study. All other are single centre studies.
C=cross-sectional, L= longitudinal, P=prospective, R=retrospective.
(ISS-insulin sensitivity score, SI-insulin sensitivity, eGDR-estimated glucose disposal rate, GIR-glucose infusion rate, ALT-alanine amino transferase, eIS-estimated insulin sensitivity, ALR-Adiponectin to Leptin ratio)

### Statistical analysis and evaluation

Meta-analysis was performed when two or more studies reported mean, standard deviation, and sample size. Metaphor package was applied for the analysis^43^. Standard Mean Difference (SMD) was calculated using R (version 4.1.1). We calculated the effect size (ES) in terms of hedges g that corrects for the sample size providing unbiased adjusted ES. Random effects model (REM) was used for quantitative meta-analysis. A forest plot was used to visualize summary of results^44^. Chi-squared test was used to measure heterogeneity (p val<0.1). The I^2^ statistic was used to estimate if the heterogeneity was considerable (I^2^>40%)^45^. The strength of relationship between parameters and traits was estimated based on the effect size (0-0.2: no effect; 0.2-0.5: small; 0.5-0.8: moderate; 0.8-1: large; >1: very large effect)^46^.

### Assessment of Sensitivity and publication bias

Funnel plots^47^ were used for visualization of publication bias. The pooled results were analysed for their sensitivity by sequential removal of individual studies and their effect on heterogeneity.

## Results

### Identification of studies for diagnostic markers of metabolic syndrome and insulin resistance

We identified 67 research articles on PubMed, 930 on SCOPUS, and 88 on Web of Science by searching keywords in titles and abstracts. After applying the filters for language and exclusion criteria, 66, 739, and 86 articles were retained. Manual search provided 3 additional studies. These articles were then imported in Rayyan^41^. In this software 78 duplicate articles were removed and 816 unique original research articles were retained. Based on the screening of abstracts and titles, 743 articles were omitted. Full text scrutiny was performed on 73 research articles, and 30 research articles were retained based on inclusion and exclusion criteria (Figure 1).

**Figure I:**
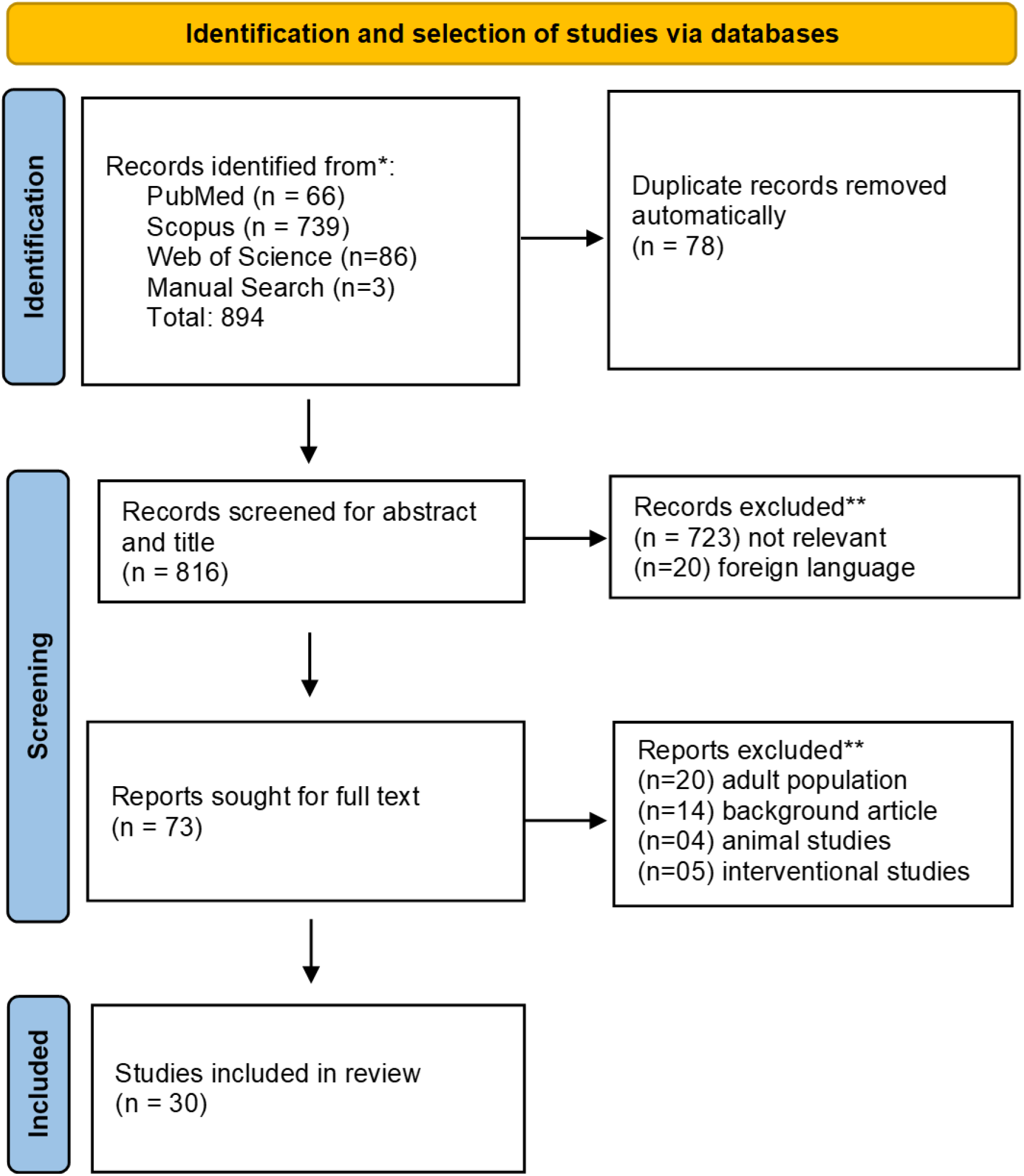
PRISMA flow diagram for the illustration of the identification and screening process. Search terms were used to compile the results in different databases and imported together in software Rayyan for duplicates removal and screening

The general nature of these research articles is mentioned in Table 3. All were observational studies with a cross-sectional or longitudinal design. The data in the studies was either prospectively collected or used retrospectively from registries and hospitals.

### Qualitative summary and characteristics of studies

As mentioned earlier, we limited our search to observational studies. There were a total of 30 studies with standard and surrogate markers of metabolic syndrome and IR in T1DM. 12 studies were based on case-control and 18 studies were cohort based. Six studies provided novel markers for IR whereas, 24 studies used existing parameters for IR and MetS. Information about ethnicity was not available for 15 studies (Table 3). Five of the 30 studies compared T1DM patients with healthy controls whereas, another 5 studies compared T1DM with T2DM patients. Four studies assessed MetS in T1DM by grouping them according to IDF criteria. The grouping of studies for IR was difficult as only two studies have classified the T1DM population on the basis of IR indices (eGDR)^48,49^.

### Assessment of markers for metabolic syndrome in T1DM

Out of 30 studies only 4 studies have grouped T1DM children as patients being metabolic syndrome positive and metabolic syndrome negative (Table 4). The parameters such as units of insulin, HbA1c, waist circumference, and lipid profile were selected for our meta-analysis. Summary statistics for fasting glucose and hypertension were not available.

**Table 4:** Studies that have categorized children with T1DM based on presence or absence of metabolic syndrome and insulin resistance.

| no | Author | Sample number | Parameters in the study | Origin/other |
| --- | --- | --- | --- | --- |
| **A. Studies based on the presence or absence of metabolic syndrome** |  |  |  |  |
| 1 | Giuliana Valerio et al., 2012<sup>67</sup> | 411 (39/372) | Insulin dose(U/kg/day), WC (cm), BMI(kg/m<sup>2</sup>), W/H ratio, HbA1c(%) | Caucasian of Italian origin, age 16-19yrs, >1yr of duration of diabetes |
| 2 | M Soliman et al., 2021<sup>57</sup> | 160 (21/139) | Duration of diabetes(yr), insulin dose(U/kg/day), HbA1C, weight, height, BMI(%), SBP(mmHg), DBP(mmHg), WC(cm), TG(mmHg), | Afrocentric ethnicity, Age<18yrs, duration of diabetes 1yr or more, |
| 3 | OY Koken et al., 2020<sup>14</sup> | 200 (21/179) | Duration of diabetes(yr), family history, insulin dose, Acanthosis, WC(cm), HbA1c(%), TG(mg/dl), HDL(mg/dl), LDL(mg/dl) | Turkish, age 13.8±2.8yrs, 4.6±3.3yrs of duration of diabetes |
| 4 | Monika Grabia et al., 2022<sup>68</sup> | 60 (20/40) | WC(cm), W/H ratio, WHtR, BMI (kg/m<sup>2</sup>), HbA1C(%), eGDR(mg/kg/min), TC(mg/dl), LDL(mg/dl), HDL(mg/dl), TG(mg/dl), SBP(mmHg), DBP(mmHg) | Polish, Age 10-17yrs, 2-7yrs of diabetes |
| **B. Studies based on the presence or absence of insulin resistance** |  |  |  |  |
| 1 | Nishtala R et al, 2020<sup>49</sup> | 175 (eGDR<7.34=58, eGDR 7.34-8.92=56, eGDR>8.93=61)) | Age, sex, ethnicity, duration of diabetes, BMI, HbA1c, eGDR, eGFR, SBP, DBP, TC, HDL, LDL, TG | Mixed(Caucasian 81.7%, African caribbean2.3%, south Asian 6.3%, other 2.3%), Age 22±1.6, 1-23yrs of diabetes |
| 2 | Dabelea et al., 2011<sup>48</sup> | 1694 (IS=1248, IR=446) | Onset age, duration of diabetes, family history, IS score, FCP, GADA titres, BMI as z score, WC | 67.9% Non-Hispanic whites (NHWs), 13.3% Hispanics, 13.4% African Americans (AAs), 4.1% Asian/ Pacific Islanders (APIs), and 1.3% American Indians (AIs), Age <20yrs, duration of diabetes is mixed population. |

In our meta-analysis using Random Effect Model (REM), WC (d =1.34, [95% CI: 0.79-1.90]) and TG (d=0.85, [95% CI: 0.14-1.55]) showed significantly large effect size whereas, HbA1C (d=0.75, [95% CI: −0.20-1.71]), and LDL (d=0.73, [95% CI: 0.15-1.32]) showed a moderate effect on MetS. The effect size was significant for LDL but not for HbA1C. On the other hand, HDL (d=-0.37, [95% CI: −0.65--0.10]) showed a significantly small negative effect. Units of insulin dose (d=0.17, [95% CI: -0.06-0.4]) also showed no significant effect on MetS (**Error! Reference source not found**.).

### Assessment of publication bias

No heterogeneity was observed for units of insulin dosage and HDL. On the other hand, HbA1c, LDL, TG, and WC showed presence of heterogeneity in datasets (Figure II). Since, the markers showed a significant heterogeneity, we decided to assess the publication bias. A funnel plot analysis was performed for all the markers mentioned above (Supplementary Fig 1). HDL and the units of insulin did not show any outliers. The publication bias was assessed for the remaining parameters which were HbA1c, WC, TG, and LDL by sequential removal of each study. The study by Monika Grabia et al 2022 strongly contributed to the heterogeneity for HbA1C, TG, and HDL. Removal of this dataset removed the heterogeneity and improved the effect size of TG (from 0.85 to 1.18) and LDL (from 0.73 to 1). The effect size of HbA1c (from 0.17 to 0.32) on the other hand, reduced. In case of WC, strong heterogeneity was contributed by the study by Soliman et al 2019. Removal of this dataset improved the effect size of waist circumference (from 1.34 to 1.63). The possible sources of heterogeneity are discussed later. In summary, the triglyceride, LDL, and waist circumference seem to have a significantly large effect on MetS (Supplementary fig 2).

**Figure II:**
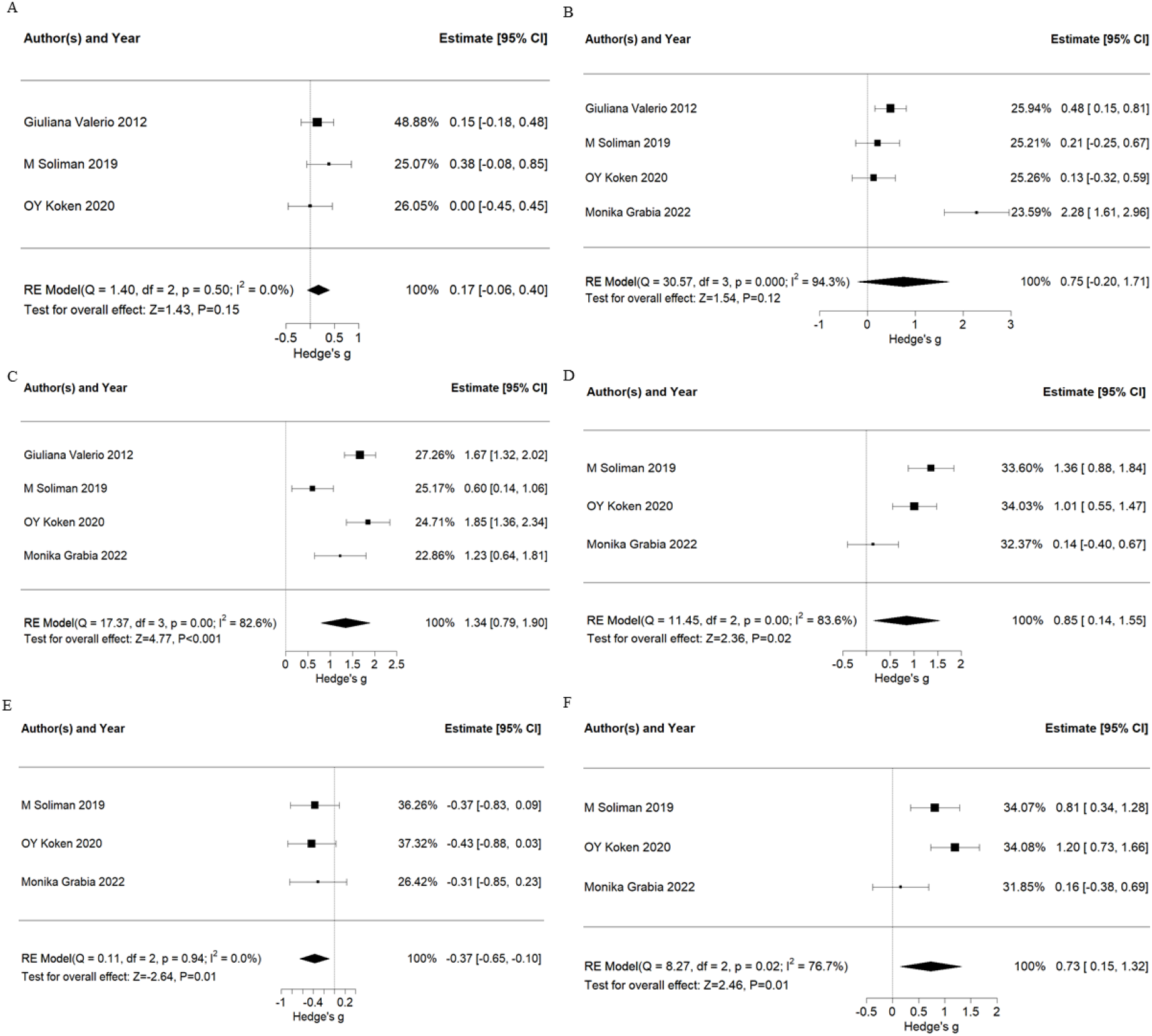
Comparison between parameters of metabolic positive and metabolic negative groups of type 1 diabetes patients. A: Insulin dosage units, B: HbA1c, C: Waist circumference, D: Triglyceride, E: High density lipoprotein, F: Low Density Lipoprotein

### Assessment of markers for IR in T1DM

Out of the 30 studies, only two studies had grouped the participants based on presence or absence of IR^48,49^. In both these studies the measurement of IR was performed by using eGDR. However, Nisthala et al., 2020, divided the patients with T1DM by eGDR_BMI_ and the association of eGDR_BMI_ with different clinical parameters was observed. The study suggested that the population in lower quartiles of eGDR_BMI_ had significantly higher levels of total cholesterol and triglycerides. Dabelea et al., 2011 attempted to segregate the population of children with T1DM and T2DM based on eGDR. The study found stronger association of IR in children with T2DM than in T1DM. The parameters to calculate eGDR and the study design were not consistent between these two studies (Table 4). Therefore, a meta-analysis could not be performed. As a result, we provide a descriptive review of other markers for IR. Some of the markers that include Volume of Oxygen uptake during peak exercise (VO2_peak_), Free Fatty Acid (FFA), Leptin, cIMT (carotid intima media thickness), have been validated using HEC. A few others markers have been validated using indices such as eGDR, SEARCH, and CACTI (Table 3).

Quantitative markers frequently used by clinicians include measurement of insulin dosage in combination with HbA1c^51^, central obesity^59^, and body fat^38^. Along with HbA1c, family history for T2DM is an important parameter. Central obesity measured by waist to height ratio (WHtR) >0.5 and BMI>95 percentile are also suggested parameters for IR^61^. Body fat estimated by thickness of triceps and subscapular skin fold have been used to predict body fat^38^. A qualitative marker: acanthosis nigricans is also used as an indicator of IR; however, it is more related to obesity than insulin resistance^48,57,58,69^.

Some of the novel quantitative markers such as adiponectin, leptin, fetuin A, and kisspeptin are being investigated for the assessment of IR. A longitudinal study in T1DM children suggested that levels of adiponectin, (a hormone produced by adipocytes with a role in insulin sensitization) were strongly related to WC and insulin dose in 20yr old adults with T1DM^60^. Adiponectin and leptin (another hormone produced by adipose tissue involved in maintenance of normal body weight), both have been studied in association with IR^53^. It has recently been suggested that leptin may act as a potential biomarker for the detection of IR in T2DM. In case of T1DM, the association of leptin with IR is not very well studied. However, a few reports suggest that fluctuations in leptin levels are observed in children and adolescents with T1DM^53,70^. Increase in fetuin A, a hepatokine and an adipokine, is associated with insulin resistance and obesity. In T1DM, this association was limited to glycemic control and as a risk predictor for complications of diabetes. Further studies to assess the role of fetuin A in insulin resistance are needed. Another hormone, Kisspeptin (produced in the hypothalamus) inversely associates with adiponectin levels and in turn, to insulin sensitivity^71^. However, the association was only studied in reproductive age female population. Further studies will be required to conclude Kisspeptin as a marker for IR. Two studies have shown an association of insulin resistance markers (increased insulin dose, increased BMI and, increased Dehydroepiandrosterone sulphate (DHEAS)) with increased micro-albuminuria^59,72^. The DHEAS is a precursor for sex hormones and is known to act as an insulin sensitizer. VO2peak which is a measure of cardiovascular and skeletal muscle oxidative function shows a significant moderate positive correlation with HEC (reduced GDR by HEC indicate IR)^50^.

Other less studied novel indicators include carbohydrate (CHO) oxidation and Delta 6 desaturase (D6D) activity. The CHO oxidation which estimates the capacity to oxidise a meal in the form of differential 13C/12C enrichment in the expired air using flow isotope mass spectrometry, has been associated with IR. The CHO oxidation showed a moderate correlation with eGDR in T1DM^73^. A high activity of D6D, a rate limiting enzyme in production of long chain Poly Unsaturated Fatty Acids (PUFA) has been associated with decreased insulin sensitivity therefore, increased activity of D6D has been suggested to be a strong marker for IR in T1DM adolescents^64^. All the novel quantitative markers are still under investigation and are not part of routine clinical applications.

## Discussion

Metabolic syndrome (MetS) and Insulin resistance (IR) in combination and independently can be the risk factors for cardiovascular diseases. Clinicians tend to use surrogate markers for the diagnosis of MetS and IR in children with T1DM. We performed a meta-analysis to investigate the effect size of these parameters for the diagnosis of MetS in paediatric T1DM patients. Inconsistency in measurement methods made it difficult to perform a similar meta-analysis for IR (Table 4).

Variations in glucose levels are expected to contribute majorly to complications in T1DM T1DM. Thus, fasting glucose is considered as a component for the diagnosis of MetS by WHO, NCEP III, and IDF (Table 1). However, mean fasting glucose levels are not provided in most studies. Waist circumference was strongly associated (with large effect size) with MetS in T1DM (Figure 1 C). In our meta-analysis, insulin dosage and HbA1c showed a poor effect size suggesting that the MetS appears independent of glycaemic condition in paediatric T1DM patients (Figure 2 A & B). Since, all four studies made use of the IDF criteria which require central obesity as a mandatory component for the assessment of MetS, this association was expected. However, this association was observed with a considerable heterogeneity. The heterogeneity was contributed by the study Soliman et al (2019). The study cohort was from Egypt and the population has been shown to have a different cut-off for waist circumference for obesity^74^. Removal of this study removed the heterogeneity and increased the effect size (Supplementary Fig 1 B). Our results fall in line with previous studies where waist circumference predicted metabolic syndrome in adults with T1DM^75^ and was significantly associated with metabolic syndrome in children who did not have diabetic^76^.

Increased triglycerides and LDL were also associated (large and moderate effect size respectively) with metabolic syndrome in patients with T1DM (Figure1 D & F). This association too was observed with heterogeneity, the source of which was identified to be the study by Monika Grabia et al (2021). Grabia et al used median and interquartile range as summary statistics whereas, others have used mean and standard deviation^42^. Though, we attempted to convert the summary statistics to means, we feel that this may have contributed to the heterogeneity in the result. The omission of this study did not alter the effect size for TG whereas, effect size for LDL improved from moderate to large (Supplementary Fig 1D). TG are already a part of IDF criteria and together with WC provide a better diagnostic efficiency for MetS^77^. The LDL also showed a strong association with MetS.

Considering that LDL is not part of the IDF criteria for MetS this association is noteworthy. Increased LDL is suggested to be a marker for cardiovascular risk. It is well documented that increased LDL levels are associated with increased CVD mortality and CVD risk^78^. Significantly increased LDL was observed in children who do not have diabetes but had predisposition to MetS^79^. Moreover, reduction in LDL levels are suggested as a treatment strategy by the IDF^80^. This reflects the significance of LDL in metabolic syndrome. However, LDL alone might be an insufficient indicator and may thus be used along with other parameters in the assessment of MetS^81^. Therefore, we suggest that increased LDL may be one of the parameters to screen for MetS in children with T1DM. However, this implementation may require further assessment with large number of studies.

HDL is one of the parameters proposed by the IDF, WHO, and NCEP III to screen MetS. HDL is known to have a negative association with metabolic syndrome which was also reflected in our analysis. All datasets showed homogeneity for HDL; however, the cumulative effect size of HDL was moderate. Other than these markers, some of the inflammatory markers such as adiponectin and Leptin are under investigation for their association with increasing cardiometabolic risk in children with MetS^66^.

For IR, we came across only two studies where young patients with T1DM were classified based on presence or absence of IR. Diverse designs and varying parameters to test insulin resistance made the compilation of studies difficult. We came across a large number of non-invasive and invasive parameters used to assess IR in T1DM. Most of them are quantitative in nature (Supplementary Table 5). Routinely used quantitative measures include BMI and waist-to-height ratio. Initially, increased BMI was one of the components for IR detection. However, with recent observations of IR in lean T1DM children^51^, it has become evident that patients especially of Asian ethnicity may follow a ‘thin fat’ phenotype with low normal BMI, and high percent fat^69^. Therefore, waist-to-height ratio may be a better marker than BMI for IR detection. Increased dose of insulin is observed in patients with IR. Insulin dosage may vary depending on the meal type, physical activity etc. Thus, insulin dose may not represent the accurate status of IR in patients with T1DM. A qualitative marker-Acanthosis Nigricans (AN) may be observed as a result of abnormal proliferation of keratinocytes due to excessive binding of insulin to insulin like growth factor receptor rather than insulin receptor^82^. Also, acanthosis is observed to be associated with obesity more than insulin resistance. Therefore, a study to assess the specificity and sensitivity of acanthosis as a marker needs to be carried out.

Among the novel markers, breath test and cIMT offer least invasive methods for detection of IR. The breath test assesses the capacity to oxidize exogenous carbohydrates which directly correlate with eGDR and ISS significantly. This is presented by enriched C12/13 in expired breath^73^. This method being non-invasive can be more applicable to large paediatric cohorts. The cIMT (carotid intima media thickness), an early sign of atherosclerosis that correlate moderately with insulin sensitivity is not a direct measure for IR and the studies using this parameter are limited to associations for assessment of cardiovascular risk in patients. The test for cIMT is expensive and would be difficult to add in to a routine check-up.

Investigations of hormones that are directly or indirectly a part of the pathogenesis of insulin resistance might help if tested. Most of these hormones are novel and under investigation. These hormones actively participate in metabolic regulation and include adiponectin, leptin, fetuin A, Kisspeptin etc. Reduced adiponectin is observed in patients with T1DM^53^. Adiponectin which is suggested to be reduced in T1DM is involved in regulation of gluconeogenesis and is an insulin sensitizer (Table 3). Adiponectin showed a good discriminatory power for IR detection in non-diabetic adolescents^83^. However, this has not been assessed specifically in patients with T1DM. Another hormone leptin that is produced by white adipose tissues shows negative correlation with insulin sensitivity. Leptin is a hormone involved in energy balance and is involved in suppression of appetite, which in turn reduces the energy intake and also increases energy expenditure. The ratio of both these hormones has been evaluated in non-diabetic adolescents^84^ however, this has not been investigated in case of children with T1DM. Fetuin A, a suggested marker for IR in non-diabetic adolescents is needed to be assessed in reference to HEC in T1DM^85^. Along with adiponectin, Kisspeptin was observed to be lower in patients with IR^71^. Kisspeptin is not very well studied; especially in case of T1DM, the role of Kisspeptin needs evaluation. An understanding of the pattern of these hormones with respect to IR provides a window for novel indices for the diagnosis of IR.

Other markers that are least understood and are under investigation include reduced D6D activity. Erythrocyte D6D activity has been suggested to be a strong marker of insulin resistance in T1DM^64^. D6D is a desaturase enzyme that introduces a double bond in a specific position of long chain fatty acids. Reduced activity of D6D can interfere with the fatty acid composition. The detailed explanation of this reduced activity is beyond the scope of our review. However, to consider D6D as an insulin resistance marker, more detailed studies are required.

### Strengths and limitations of the study

To the best of our knowledge, this is the first systematic review and meta-analysis for assessment of surrogate markers for metabolic syndrome and a systematic review for insulin resistance in children with T1DM. However, for the insulin resistance, the studies are reported in different forms of indices which made it difficult for us to compile them for the assessment of IR markers. Also, this systematic review could not assess the effect of age and pubertal status on the accuracy of markers of metabolic syndrome and IR. The number of studies available for meta-analysis are very small hence, with increasing reports there are chances that the results may improve in future.

## Conclusion

From the results it can be concluded that in the paediatric population with T1DM, markers of glycaemic control are not associated with MetS. Other than TG and HDL, LDL may also be considered in the diagnostic criteria for MetS. A combination of WC and TG may increase the efficacy of MetS diagnosis in paediatric patients with T1DM. Many novel markers currently under investigation for the diagnosis of IR need evaluation against HEC. These markers may be used in combination to increase the accuracy of IR diagnosis.

## Data Availability

All data produced in the present work are contained in the manuscript

https://github.com/sukeshini321khandagale/Meta_analysis_Script

## Declaration Section

### Author’s Contribution

SBK and VK performed the systematic literature search. SBK performed the statistical analysis. SBK and SPK wrote the manuscript. AK and SPK contributed to conceptual design of the study.

### Funding

This research received no specific grant from any funding agency in the public, commercial, or not-for-profit sectors.

### Ethics declaration

No ethical approval was needed as the data was collected from previous published studies in which the informed consent was obtained by primary investigators.

### Competing interests

The authors declare no competing interest

## Acknowledgements

SBK and VK thank SIU for research fellowships. SPK is a beneficiary of an extramural funding from SERB (SRG/2020/001414).

## Notes

### Competing Interest Statement

The authors have declared no competing interest.

### Clinical Protocols

https://www.crd.york.ac.uk/prospero/CRD42023418954

### Funding Statement

This study did not receive any funding

### Summary of Updates

Supplementary file added

