## Supplementary Figure for "Surrogate markers of metabolic syndrome (MetS) and insulin resistance (IR) in children and young adults with type 1 diabetes: A systematic review & meta-analysis"

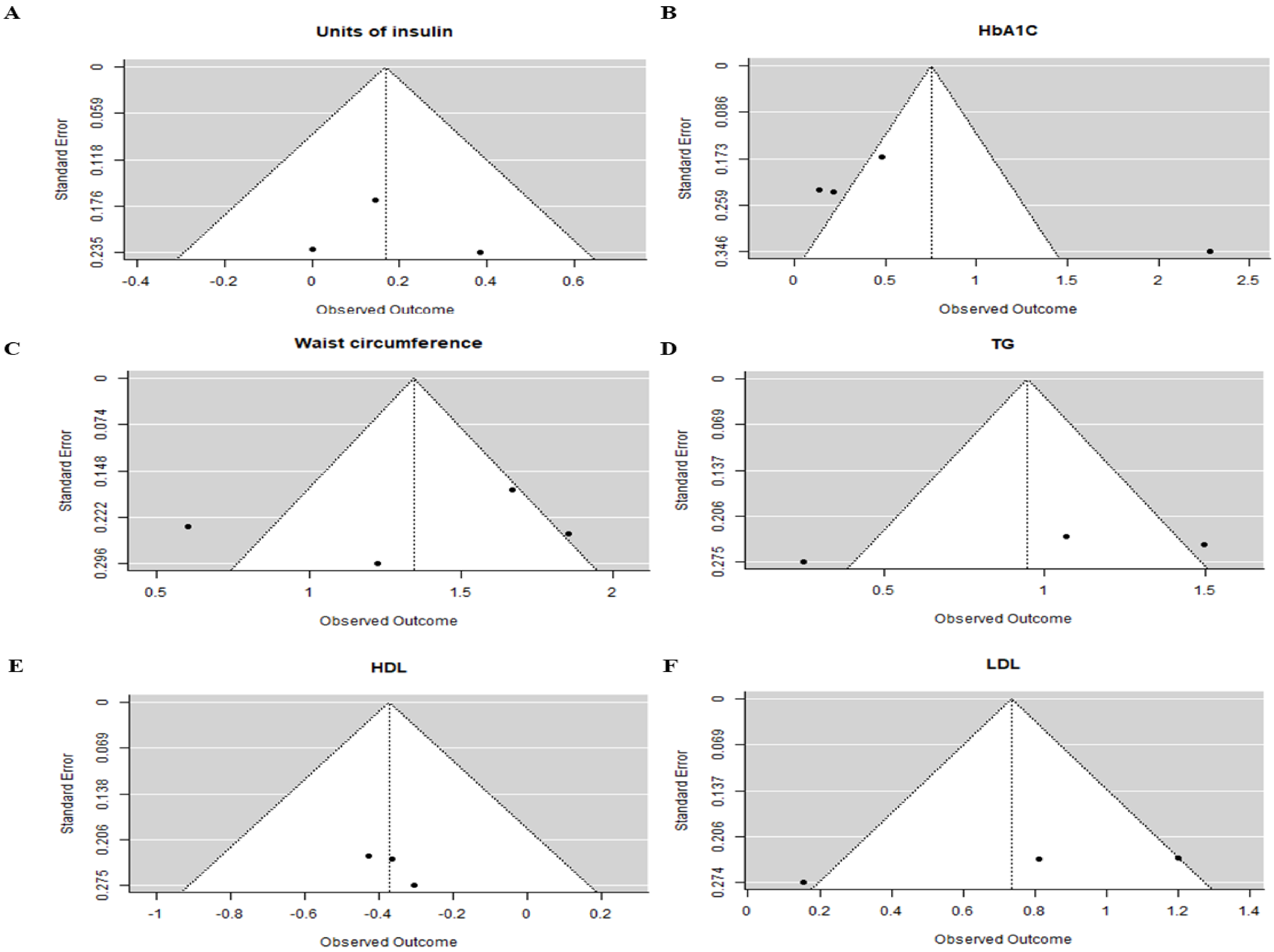


Figure 1: Funnel plots for all parameters representation of asymmetry between studies.

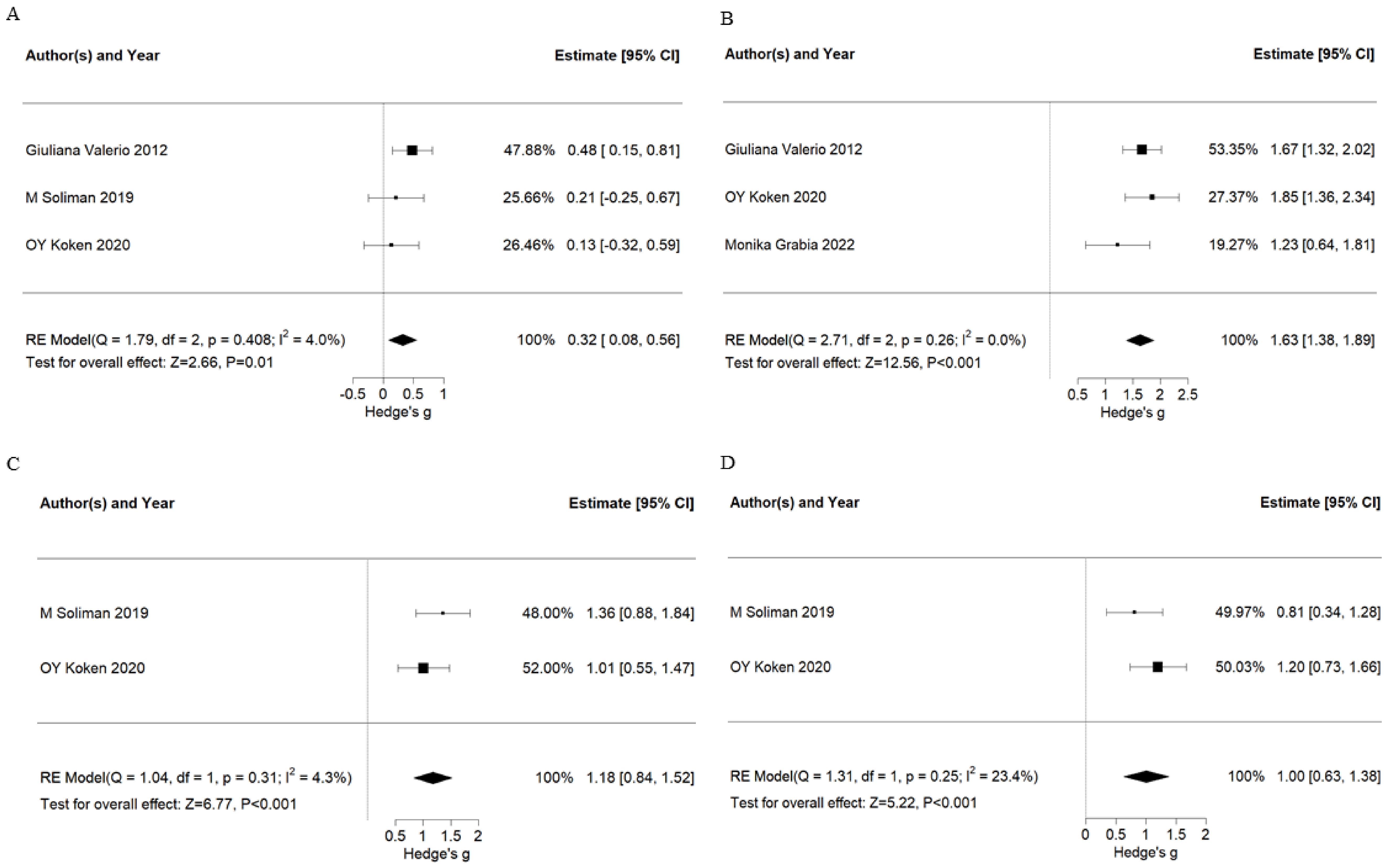


Figure 2: Forest plots after correction for heterogeneity. (A)HbA1C (B)WC (C) TG (D)LDL. Monika grabia is an outlier for multiple studies. This may be because the values are given in median and IQR that we converted to mean and median. These plots are after the removal of heterogeneity.
