## Supplementary Table for "Surrogate markers of metabolic syndrome (MetS) and insulin resistance (IR) in children and young adults with type 1 diabetes: A systematic review & meta-analysis"

**Supplementary Information**

Authors: Sukeshini Khandagale^1^, Vinesh Kamble^1^, Chirantap Oza ^2^, Shital Bhor^2^, Anuradha Khadilkar^2^#, Satyajeet Khare^1^#

1. Symbiosis School of Biological Sciences, Symbiosis International University, Pune 412115
2. Hirabai Cowasji Jehangir Medical Research Institute, Jehangir Hospital, Pune 411001

**Corresponding authors:**

Satyajeet Khare

Anuradha Khadilkar

Supplementary Table 1: eGDR indices provided by different studies

|  | **Provided by** | **Equation** | **Cut-off** | **Population type/size/ethnicity** |
| --- | --- | --- | --- | --- |
| 1 | Williams et al 2000^40^ (Pittsburg EDC) | eGDR = 24.31-12.22×(WHR)-3.29×(Hypertention)-0.57×(HbA1[%]) | Does not provide cutoff, usually studied by dividing groups in tertiles or quartiles^37^. Lower the eGDR, higher IR | Adult T1DM (compared with HEC) |
| 2 | Kilpatrick et al^36^ (DCCT-EDIC cohort) (2007) | eIS=24.31-(12.22*WHR) -(3.29*Hypertension)-(0.57*A1C), where the units are mg^-1^.kg^-1^.Min^-1^. | Does not provide cutoff (modified version of eGDR using HbA1C rather than HbA1. Lower the eGDR, higher IR | Adult T1DM (modified version) |
| 3 | Texeiria et al 2014^37^ (SEARCH) | eGDR =24.4-(12.97 ×W/H)- (3.39×AH)-(0.60×A1c)  LogeIS = 4.64725 - 0.02032 (W, cm) - 0.0977 (A1c, %) - 0.00235 (TG, mg/dl) | ≤6.2ml/kg/min for IR | Adult T1DM |
| 3 | Dabelea et ala 2011^34^ (SEARCH) | IS scores = Exp(4.64725-0.02032(waist[cm])-0.09779(HbA1c[%])-0.00235(TG[mg/dlL]) | Do not have cuttoff, but shows correlation with clinical parameter | Adolescence with T1DM, T2DM and non-diabetic (compared with HEC) |
| 4 | Snell-Burgeon et al 2011^39^ (CACTI) | eIS = Exp(4.1075-0.01299×(waist[cm])-1.05819×(insulin dose)-0.00354×(TG[mg/dL)-0.00802×(DBP[mmHg]) | Do not have cuttoff, parameters tested as models with and without adiponectin and fasting/non-fasting state | Adult T1DM (compared with HEC) |
| 5 | Zheng et al 2017^86^ | lnGDR= 4.964 -0.121*HbA1c(%)-0.012*DBP(mmHg)-1.409*WHR | No cut-off | Adult T1DM |
| 6 | Uruska et al 2018 | TG/HDL-C and VAI  (for women = [waist circumference/36.58+(1.89xBMI)]x(TG/0.81)x(1.52/HDL) and for men = [waist circumference/39.68+(1.88xBMI)]x(TG/1.03)x(1.31/HDL)) | GDR(by kilpatric)<4mg/kg/min are IR. | Adult T1DM |
| 7 | Cano et al., 2020^87^ | Equation provided by Kiplpatrick^36^ et al and Duca^35^ et al | eIS-EDC <8.08mg/kg/min  eIS-CACTI 3.43mg/kg/min (both for high risk of CVD) | Adult with T1DM (18-65yrs) |
| 8 | Hermosillo et al 2020 | Equation provided by Kilpatrick^36^ | eGDR<7.32mg/kg/min (for MS in patients with T1DM) | Adults with T1DM (>18yrs) |
| 9 | Oza et al., 2022 | SEARCH equation | IS<5.485 | Age 12-18yrs |

Supplementary Table 2: Categorization of insulin resistance markers

|  | Qualitative | Quantitative |
| --- | --- | --- |
| Invasive | NA | HEC, Adiponectin levels, Leptin levels, Fetuin A levels, Kisspeptin levels, Erythrocyte D6D activity, Units of insulin |
| Non invasive | Breath test, cIMT, Acanthosis nigricans | Waist circumference (cm), BMI (kg/m^2^), total body fat, thickness of tricep, subscapular skinfold measurement |
| Combination | Indices provided by EDC, DCCT-EDIC, SEARCH, CACTI | |

**Note:** HTN(Hypertension), cIMT (carotid intima media thickness), D6D (Delta 6 desaturase)
